# Understanding and addressing challenges for Advance Care Planning in the COVID-19 pandemic: An analysis of the UK CovPall survey data from specialist palliative care services

**DOI:** 10.1101/2020.10.28.20200725

**Authors:** A Bradshaw, L. Dunleavy, C. Walshe, N. Preston, R. Cripps, M.B. Hocaoglu, S. Bajwah, M. Maddocks, A. Oluyase, K.E. Sleeman, I.J. Higginson, L.K. Fraser, F.E.M Murtagh, On behalf of the CovPall study team

## Abstract

**Background:** During the COVID-19 pandemic, specialist palliative care services have an important role to play conducting high-quality and individualised Advance Care Planning discussions. Little is known about the challenges to Advance Care Planning in this context, or the changes services have made in adapting to them.

**Aim:** To describe the challenges experienced, and changes made to support, Advance Care Planning at the height of the COVID-19 pandemic.

**Design:** Cross-sectional on-line survey of UK palliative and hospice services’ response to COVID-19. Closed-ended responses are reported descriptively. Open-ended responses were analysed using a thematic Framework approach.

**Respondents:** 277 UK palliative and hospice care services.

**Results:** 37.9% of services provided more Advance Care Planning directly. 58.5% provided more support to others. Some challenges to Advance Care Planning pre-dated the pandemic, whilst other were COVID-19 specific or exacerbated by COVID-19. Six themes demonstrated challenges at different levels of the Social Ecological Model, including: complex decision making in the face of a new disease; maintaining a personalised approach; COVID-specific communication difficulties; workload and pressure; sharing information; and national context of fear and uncertainty. Two themes demonstrate changes made to support Advance Care Planning, including: adapting local processes and adapting local structures.

**Conclusions:** Professionals and healthcare providers need to ensure Advance Care Planning is individualised by tailoring it to the values, priorities, and ethnic, cultural, and religious context of each person. Policymakers need to consider carefully how high-quality, person-centred Advance Care Planning can be resourced as a part of standard healthcare ahead of future pandemic waves.

**Key Statements:** *What is already known about the topic?:*

– An important part of palliative care’s response to COVID-19 is ensuring that Advance Care Planning discussions occur with patients and their care networks
– High quality Advance Care Planning is viewed as a process that adopts a holistic, collaborative, and individualised approach
– Prior to COVID-19, challenges to Advance Care Planning included time constraints, lack of training, fears of taking away hope, limited resources, and insufficient knowledge

*What this paper adds?:*

– The COVID-19 pandemic exacerbated already-existing challenges to conducting high-quality, individualised Advance Care Planning, including the ability to maintain a personalised approach and sharing information between services
– COVID-specific challenges to Advance Care Planning exist, including the complexities of decision-making for a novel disease, communication issues, and workload pressures
– In responding to these challenges, services adapted local processes (prioritising specific components, normalisation and integration into everyday practice) and structures (using technology, shifting resources, collaboration) of care

*Implications for practice, theory or policy:*

– COVID-19 has provided an opportunity to re-think Advance Care Planning in which the starting point to any discussion is always the values and priorities of patients themselves
– Providers and policymakers need to urgently consider how high-quality Advance Care Planning can be resourced and normalised as a part of standard care across the health sector, ahead of future or recurrent pandemic waves and in routine care more generally
– We provide questions for health professionals, services, and policy makers to consider in working towards this

## Introduction

In March 2020, the World Health Organisation declared Coronavirus (COVID-19) a global pandemic, with an estimated global mortality rate of 3.4%, increasing with age and co-morbidities.^(1)^ The number of patients suffering and dying from COVID-19-related illness is placing huge pressure on healthcare systems across the world.^(2)^ To date, there have been 29,155,581 confirmed cases and 926,544 deaths attributable to COVID-19. ^(3)^

Worldwide, specialist palliative care services have an important role to play in responding to the pandemic and are skilled in delivering person-centred symptom control and making complex decisions in the face of uncertainty. ^(2, 4, 5)^ One crucial aspect of decision making in palliative care - and even more so within the context of the pandemic - is ensuring that Advance Care Planning (ACP) discussions occur with patients (and their families).

Adapting existing person-centred definitions ^(6, 7)^, we define high quality ACP as ‘considerations and activities to best prepare for future care, including: identifying values based on past experiences and quality of life; choosing proxy decision-makers wisely and verifying that they understand their role; deciding whether to grant leeway (and how much) in proxy decision making, and; informing other family of wishes in advance to reduce or prevent conflict’. As a person’s preferences and priorities are complex and may change over time, ^(8, 9)^ we view ACP as a process, not a one-time event or document. ^(10)^ In the COVID-19 pandemic, it is crucial that healthcare professionals have high quality and timely ACP discussions with patients and their families, to enhance the likelihood of improved outcomes and satisfaction.^(11-13)^ However, this presents multiple challenges.

Patient (unpredictable disease/prognosis, insufficient knowledge of health status, anxiety, and denial), ^(13, 14)^ professional (time constraints, lack of communication training/skills, fears of taking away hope), ^(13-15)^ and system-wide (limited resources and unclear responsibilities) ^(14-16)^ challenges exist to initiating and following-up ACP discussions. Currently, however, there is lack of evidence regarding how these issues manifest during the COVID-19 pandemic, or what may be done to address these challenges. Addressing these issues is crucial in optimising the specialist palliative care response to the COVID-19 pandemic and for adapting to future increases in the need for palliative care.^(17, 18)^ This study aims to describe the challenges that UK palliative care services experienced regarding ACP at the height of the COVID-19 pandemic and the changes made to support ACP.

## Methodology and Methods

### Design and participants

The CovPall study is a rapid multinational observational study of palliative care during COVID-19. This paper reports the cross-sectional on-line survey of hospice and specialist palliative services in the UK given that understandings of ACP during COVID-19 are dependent on the cultural and policy contexts in which they are conducted. Services providing hospice and specialist palliative care across inpatient palliative care, hospital palliative care, home palliative care, and home nursing settings were eligible for participation and recruited through palliative care and hospice organisations (Sue Ryder, Hospice UK, Marie Curie, European Association of Palliative Care, Together for Short Lives, and the palliativedrugs.com and www.pos-pal.org network) between April and July 2020. We invited service leads to complete the survey online with the aim of receiving responses from all hospice and specialist palliative care services.

Ethical approval was obtained from King’s College London Research Ethics committee (LRS-19/20-18541). The CovPall protocol is registered (ISRCTN16561225) and is reported according to STROBE ^(19)^ and CHERRIES checklists. ^(20)^

### Survey and data collection

REDCap was used to securely build and host the survey which aimed to understand how palliative care services responded to the COVID-19 pandemic, and comprised of closed- and free-text responses (full survey in supplementary file 1 and procedures for survey are provided in supplementary file 2). This paper focuses on the impact of COVID-19 on ACP via analyses of two closed-ended and two free-text questions (see Table 1).

**Table 1:** Closed and free-text survey questions analysed in this study.

|  |  |  |
| --- | --- | --- |
| **Closed questions** | 4.13. Would you say you are now involved directly with patients/families in advance care planning?<br>4.14. Would you say you are now involved advising/supporting others and/or educating about advance care planning? | 1. A lot more<br>2. Slightly more<br>3. About the same<br>4. Slightly less<br>5. Much less |
| **Open questions** | 4.15. In what ways (if any) have you changed how you are supporting advance care planning?<br>4.16. What would you say are the main challenges for advance care planning during the COVID-19 pandemic? | Free text response |

### Data analysis

Anonymised quantitative data items were summarised descriptively. Free text responses were analysed in NVivo (v12) using a thematic Framework approach. ^(21)^ This allowed within- and between-case analyses to be made to explore how key contextual variables related to main themes. The following iterative steps were followed: (1) familiarisation and coding; (2) developing an analytic framework; (3) indexing; (4) charting (by developing matrices to understand differences in main themes across key variables); and (5) interpreting the data. During the development of our analytic framework, we recognised that responses to the challenges to ACP free-text question could be understood through using an adapted version of the Social Ecological Model. ^(22, 23)^ This model recognises that challenges to ACP exist at multiple interacting levels (individual, interpersonal, within teams/services, between teams/services, and national).

We adopted a relativist approach to rigour ^(24)^ by using lists of criteria on what researchers agree constitutes high quality qualitative analysis ^(25-27)^ as a starting point and then selecting criteria appropriate to the context, purposes, and methodology of this study (table 2).

**Table 2:**
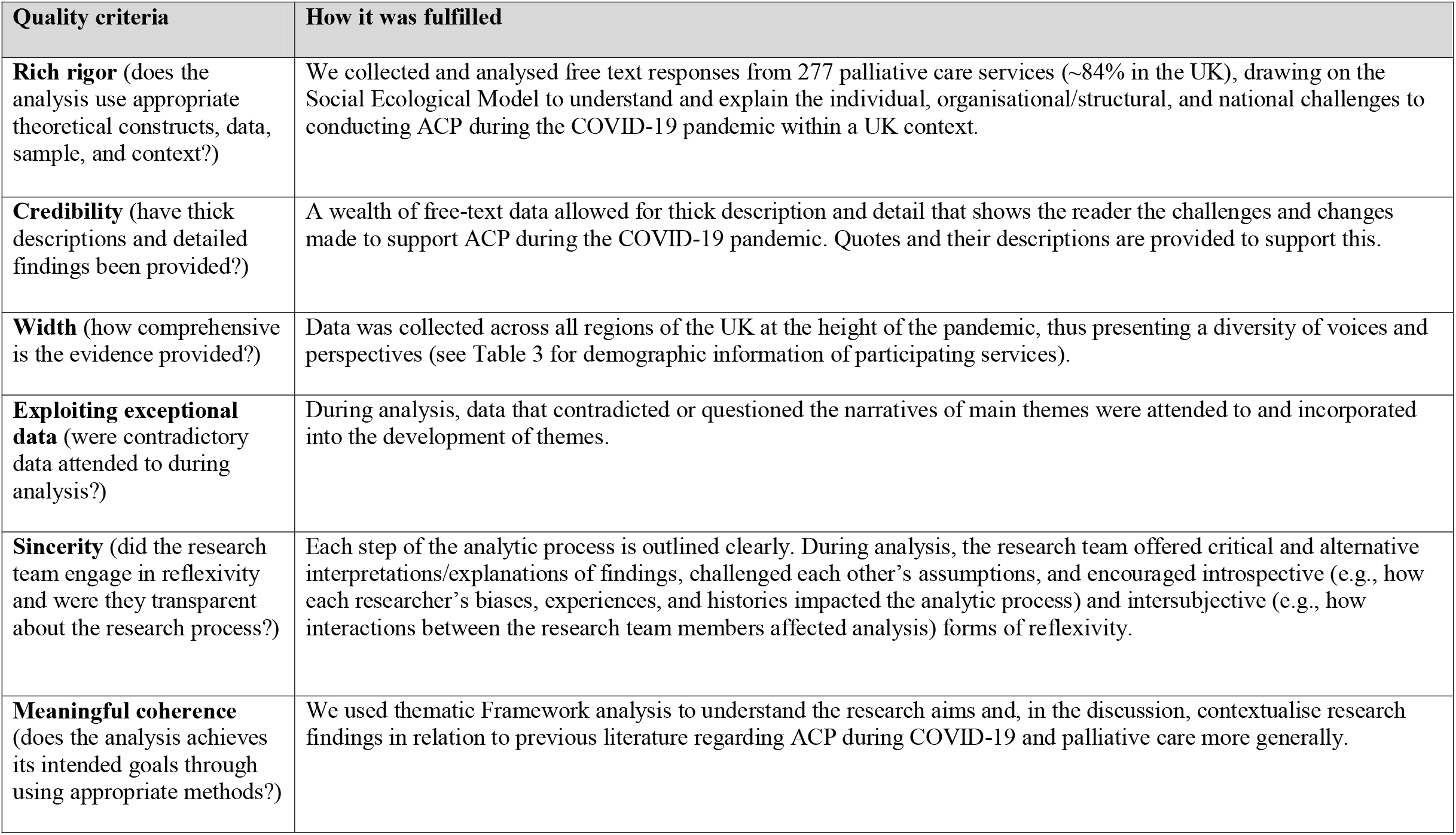
List of quality criteria selected for ensuring a rigorous qualitative analysis and how it was fulfilled in this study.

| Quality criteria | How it was fulfilled |
| --- | --- |
| **Rich rigor** (does the analysis use appropriate theoretical constructs, data, sample, and context?) | We collected and analysed free text responses from 277 palliative care services (~84% in the UK), drawing on the Social Ecological Model to understand and explain the individual, organisational/structural, and national challenges to conducting ACP during the COVID-19 pandemic within a UK context. |
| **Credibility** (have thick descriptions and detailed findings been provided?) | A wealth of free-text data allowed for thick description and detail that shows the reader the challenges and changes made to support ACP during the COVID-19 pandemic. Quotes and their descriptions are provided to support this. |
| **Width** (how comprehensive is the evidence provided?) | Data was collected across all regions of the UK at the height of the pandemic, thus presenting a diversity of voices and perspectives (see Table 3 for demographic information of participating services). |
| **Exploiting exceptional data** (were contradictory data attended to during analysis?) | During analysis, data that contradicted or questioned the narratives of main themes were attended to and incorporated into the development of themes. |
| **Sincerity** (did the research team engage in reflexivity and were they transparent about the research process?) | Each step of the analytic process is outlined clearly. During analysis, the research team offered critical and alternative interpretations/explanations of findings, challenged each other's assumptions, and encouraged introspective (e.g., how each researcher's biases, experiences, and histories impacted the analytic process) and intersubjective (e.g., how interactions between the research team members affected analysis) forms of reflexivity. |
| **Meaningful coherence** (does the analysis achieves its intended goals through using appropriate methods?) | We used thematic Framework analysis to understand the research aims and, in the discussion, contextualise research findings in relation to previous literature regarding ACP during COVID-19 and palliative care more generally. |

## Findings

### Characteristics of survey sample and ACP provision

277 UK palliative care services responded (see Table 3). 248 services reported caring for patients with suspected or confirmed COVID-19, and 16 services reported no suspected or confirmed cases of COVID patients (13 missing data). The number of COVID-19 patients seen ranged from 0-400, (median 14; IQR 5-52). 37.9% of responding services were directly providing more ACP and 58.5% were providing more advice to others about ACP. The vast majority (92.4%) of those who were providing more direct ACP were also providing more advice to others about the ACP process.

**Table 3:**
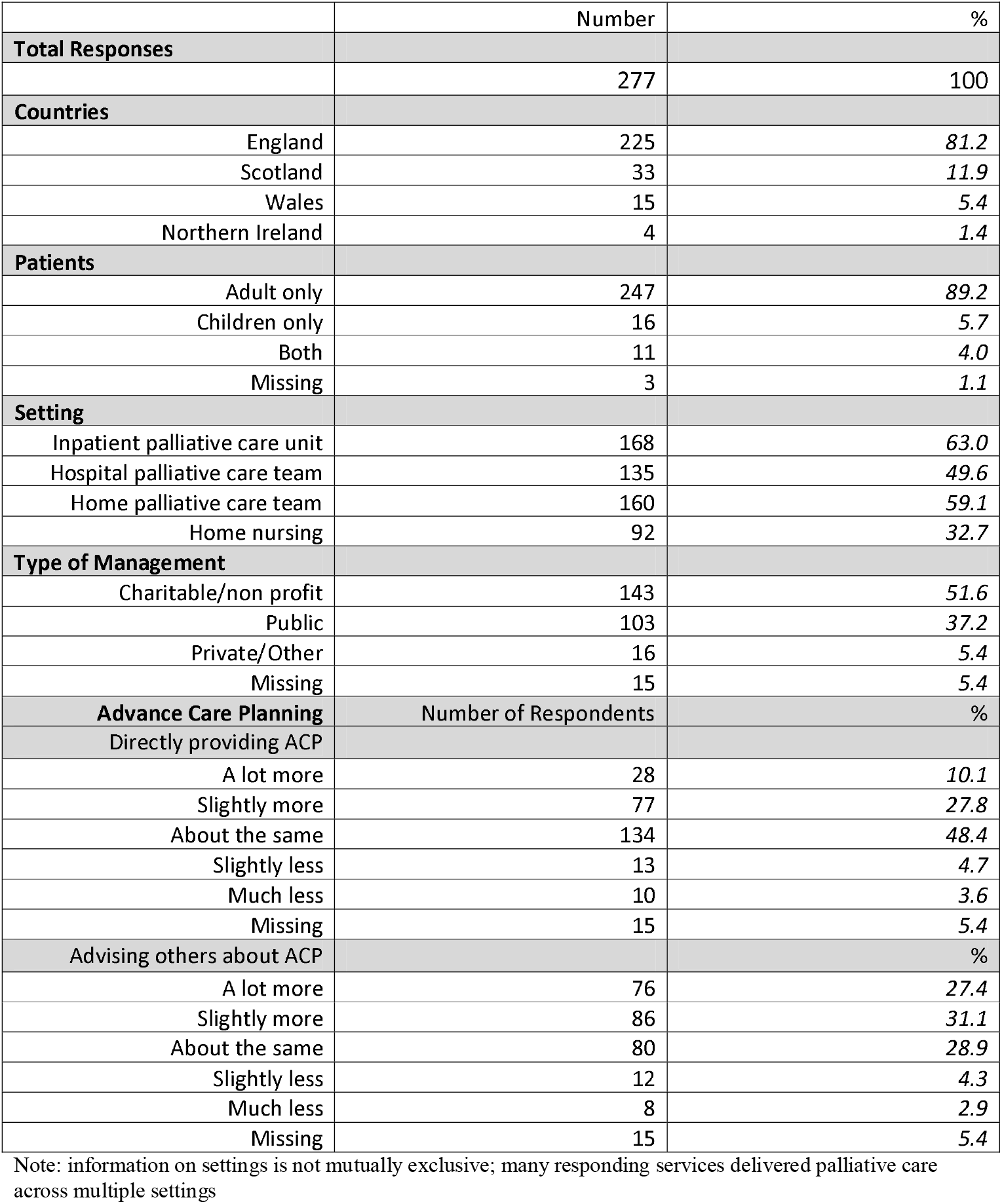
Demographic information of participating palliative care services and an overview of UK participants’ responses to closed-ended CovPall survey questions on Advance Care Planning.

|  | Number | % |
| --- | --- | --- |
| **Total Responses** |  |  |
|  | 277 | 100 |
| **Countries** |  |  |
| England | 225 | 81.2 |
| Scotland | 33 | 11.9 |
| Wales | 15 | 5.4 |
| Northern Ireland | 4 | 1.4 |
| **Patients** |  |  |
| Adult only | 247 | 89.2 |
| Children only | 16 | 5.7 |
| Both | 11 | 4.0 |
| Missing | 3 | 1.1 |
| **Setting** |  |  |
| Inpatient palliative care unit | 168 | 63.0 |
| Hospital palliative care team | 135 | 49.6 |
| Home palliative care team | 160 | 59.1 |
| Home nursing | 92 | 32.7 |
| **Type of Management** |  |  |
| Charitable/non profit | 143 | 51.6 |
| Public | 103 | 37.2 |
| Private/Other | 16 | 5.4 |
| Missing | 15 | 5.4 |
| **Advance Care Planning** | **Number of Respondents** | **%** |
| **Directly providing ACP** |  |  |
| A lot more | 28 | 10.1 |
| Slightly more | 77 | 27.8 |
| About the same | 134 | 48.4 |
| Slightly less | 13 | 4.7 |
| Much less | 10 | 3.6 |
| Missing | 15 | 5.4 |
| **Advising others about ACP** |  | **%** |
| A lot more | 76 | 27.4 |
| Slightly more | 86 | 31.1 |
| About the same | 80 | 28.9 |
| Slightly less | 12 | 4.3 |
| Much less | 8 | 2.9 |
| Missing | 15 | 5.4 |

Note: information on settings is not mutually exclusive; many responding services delivered palliative care across multiple settings

### Free text responses

#### Section 1: Challenges to ACP

Six themes and two sub-themes represent the challenges to ACP and how these were understood through the different levels of the Social Ecological Model. Whilst some of these challenges were specific to COVID-19, others were general challenges exacerbated by the pandemic (see Figure 1).

**Figure 1:**
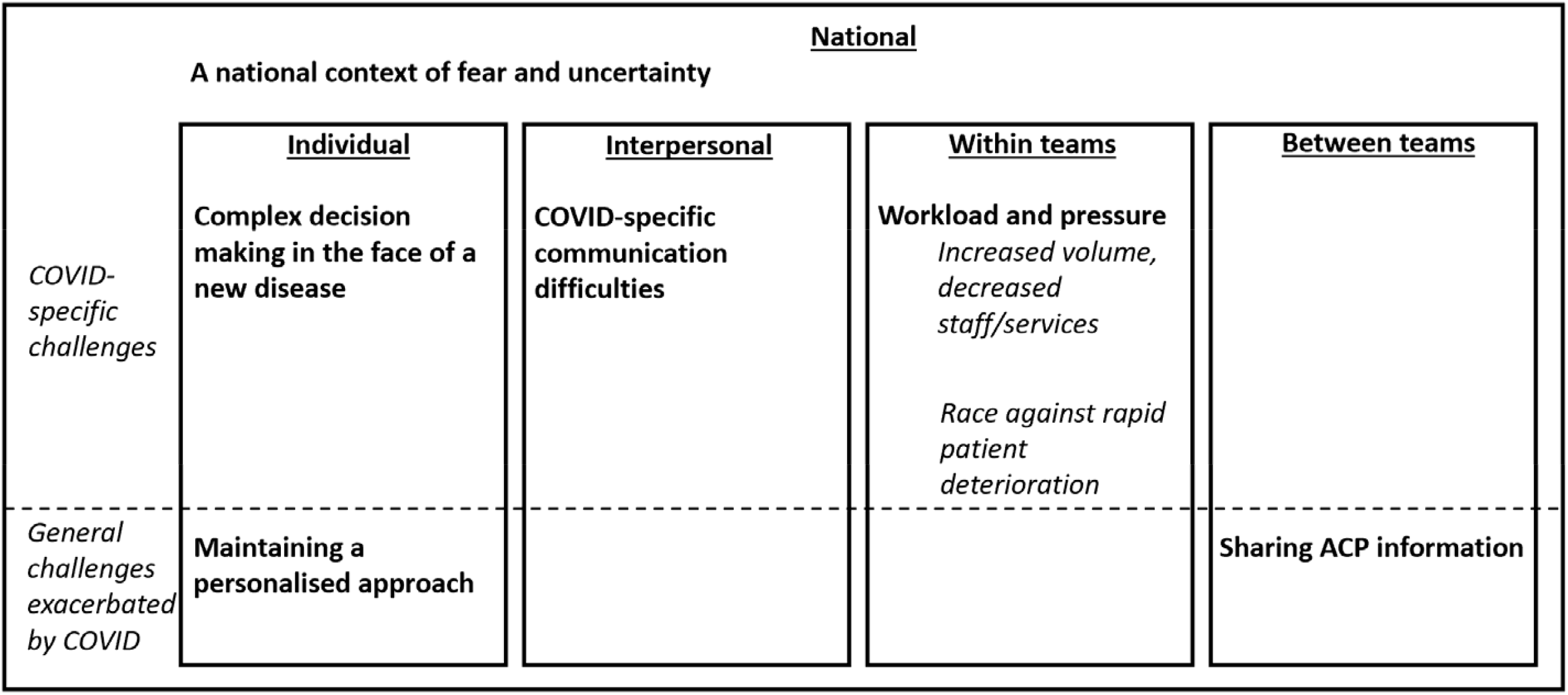
An overview of the **themes** and *sub-themes* that represent the challenges to ACP in the context of COVID-19, and how they relate to the different levels of the Social Ecological Model. Note: This diagram is representative of findings related to section 1 only (the challenges to ACP)

#### National Level

##### Theme 1: A national context of fear and uncertainty

ACP discussions were challenging because they took place in a national context of fear and uncertainty. Fears across society – alongside national policies on social/physical distancing – provided a contextual backdrop through which challenges at other levels of the Social Ecological Model may be understood.

A major source of fear and uncertainty was that many patients, their families, and healthcare professionals perceived that clinical decisions were being made on the basis of limited resources, rationing of treatments and services, and the government policy to ‘protect the NHS’. There was particular concern that people who were older, had comorbidities, were disabled, or from Black, Asian and Minority Ethnic backgrounds were more likely to be discriminated through the adoption of a blanket– as opposed to a person-centred - approach to decision-making:

> *‘Perception in public that [ACP] is about rationing rather than sensible clinical judgement. In young adult/transition work, huge fear among disabled communities and the perception that they will be denied potentially effective interventions due to discrimination’*. [Hospital Palliative Care Team, Adult Service, Scotland]
>
> *‘Family and patient concerns around ‘blanket’ and CPR [cardiopulmonary resuscitation] decisions’*. [Hospital Palliative Care Team, Adult Service, England]

Respondents reported that media coverage – regardless of its accuracy - on issues such as blanket/generalised decisions, rationing of treatments, and the role (and limits of) ventilatory support, exacerbated the aforementioned public fears and uncertainties:

> *‘My views on advance care planning remain the same as pre-COVID; it should be individualised to improve patient care. I have continued to practice in this way. The media has covered how during the pandemic there have been some cases when the way it has been delivered has led to those at the receiving end feeling as though their focus has been on protecting services as opposed to the individual.’* [Multiple Settings, Adult Service, Scotland]

#### Individual level

##### Theme 2: Complex decision-making in the face of a new disease

The rapid onset of a novel disease with so many uncertainties meant that decision-making during ACP became even more complex and challenging. Uncertainties regarding the clinical trajectory and prognosis of COVID patients contributed to the challenges of ACP because COVID seemed to affect people in different ways; recovery, mortality, and outcomes varied between patients making it difficult to use past experience to inform subsequent decisions. Moreover, profound uncertainties of a different order existed that were related to knowing nothing about COVID-19 (e.g., its death/infection rate, or whether it was acute/chronic, etc.):

> *‘The uncertainty of response. Patients with advanced disease have survived while those with no underlying medical conditions have died. The ability to know the course of the illness, and make informed decisions with patients in light of that uncertainty, is challenging. It requires a dynamic approach to decision making which is difficult to sensitively achieve at times of high stress in medical systems.’* [Multiple Settings, Adult Service, England]

One aspect of decision making that was particularly complex and challenging was surrounding service provision and treatment options. This included discussing what services and treatments were appropriate/available, predicting how patients may respond to treatments, treatment limitations, and how any decisions on these issues were subject to dynamic changes in a person’s health status:

> *‘Uncertainty about treatment availability, potential prognosis on an individual level, when to stop medical interventions like CPAP [meaning continuous positive airway pressure ventilation]/high flow oxygen’*. [Hospital Palliative Care Team, Adult Service, England]
>
> *‘Some of the decisions about limitation of treatment may be appropriate while the patient has COVID but may not be if they recover and then experience different health conditions. I wonder if this review process is happening’*. [Hospital Palliative Care Team, Adult Service, England]

##### Theme 3: Maintaining a personalised approach

Respondents reflected on how the abruptness of the pandemic made it difficult to avoid ACP becoming a ‘tick-box’ exercise in which generalised decisions were made:

> *[One main challenge was reported as] ‘avoiding ACP becoming part of a tick box culture and remaining a meaningful conversation about what is important to a patient, ensuring the promotion of ACP is for the benefit of the patient and not motivated by limited resources.’* [Multiple settings, Adult Service, Scotland]

A prominent challenge to maintaining an individualised approach – particularly with regards to preferred place of care/death - was that ACP discussion were occurring in the context of limited choices regarding discharge options. This was either because some services refused to accept COVID patients or because there was a reluctance in being discharged to settings where there were COVID positive patients and consequent visiting restrictions:

> *‘Care options are different - not able to access care homes or the hospice as preferred place of care/death, especially in the first 5 weeks’*. [Home Palliative Care Team, Adult Service, Wales]

#### Interpersonal Level

##### Theme 4: COVID-specific communication difficulties

Policies on physical/social distancing and the use of personal protective equipment presented COVID-specific communication challenges. A common communication challenge reported was having remote and telephone ACP conversations. Lack of face-to-face contact meant that many healthcare professionals felt that they had lost some of the ‘tools’ that were integral to their practice during these exchanges:

> *‘The reduced face-to-face contact and social distancing feels like we have had our tools taken away from us - emphasizing the importance of advanced communication skills - listening and responding appropriately, ensuring clear understandable language… The ward teams have needed to give bad news over telephone contact which is not usual practice - advance care planning over the telephone is markedly harder than it is face-to-face.’* [Hospital Palliative Care Team, Adult Service, England]

These lost ‘tools’ included the ability to draw on non-verbal clues (e.g., physical touch, reading the environment/patient cues), and develop trusting/respectful relationships prior to conversations; things deemed fundamental in managing the sensitivities, compassion, and nuances of ACP conversations:

> *‘Not being able to have face-to-face discussions when having sensitive conversations, not being able to physically touch patients and their loved ones who may crave physical comfort such as a hug or hand being held.’* [Multiple Settings, Adult Service, England]
>
> *‘Staff finding it difficult to have those conversations with people who they haven’t met before and having to do it remotely feels impersonal and harsh.’* [Multiple Settings, Adult Service, England]

Even if face-to-face discussions were possible, personal protective equipment acted as a physical barrier which made it difficult to use non-verbal communication to display compassion/empathy or provide physical forms of comfort:

> *‘Personal protective equipment has been a challenge as difficult to see facial expressions or comfort a family member during difficult distressing discussions.’* [Multiple Settings, Adult and Children Services, England]

Regardless of whether discussions were remote or face-to-face, an overarching challenge to communicating ACPs during the pandemic was the difficulty of involving families in conversations.

> *‘The restrictions on visiting make it more difficult to involve families and often the family haven’t seen the patient for some time and don’t have that visual cue of how unwell they are.’* [Hospital Palliative Care Team, Adult Service, England]

There were concerns that people from ethnic minority backgrounds may have been disproportionately affected by these communication difficulties:

> *‘it is much harder in those patients / families that you haven’t seen face to face, and particularly when there are cultural or language barriers or capacity issues preventing a conversation with the patient.’* [Specialist Palliative Home Care Service, Adult Service, England]

#### Within teams and services level

##### Theme 5: Workload and pressure

###### Sub-theme 1: Increased volume, decreased staff and services

The increase in the number of patients who needed ACP discussions (for new referrals and reviewing patients already on their caseload) meant that teams had to work longer, harder, and quicker to ensure that timely ACP discussions occurred:

> *‘Volume of people who need them.’* [Multiple Settings, Adult Services, England] *‘The numbers involved, particularly [in the] care home sector.’* [Multiple Settings, Adult Service, Northern Ireland]

Exacerbating this was a decrease in the availability of staff (due to absence, deployment to other services, and understaffing):

*‘Staff availability for distribution. Reaching all required professionals, some of the other professionals are working differently so may not be as available, also potential increase in staff absence may present a challenge.’* [Multiple Settings, Children Service, England] *‘More difficult to conduct as not seeing patients earlier in their prognosis as no day care facilities.’* [Multiple Settings, Adult Service, England]

###### Sub-theme 2: A race against rapid patient deterioration

Compounding an increased workload and pressure was the rapid clinical deterioration of COVID patients which resulted in a perpetual race against time to engage in discussions before they became too ill, lost capacity, or died:

> *‘There wasn’t time for advance care planning with patients with COVID - prognosis was sudden and very short.’* [Hospital Palliative Care Team, Adult Service, England]
>
> *‘The hospital palliative care team have had an increase in referrals of very unwell semiconscious/unconscious with severe respiratory failure and high O2 requirements who are imminently dying and too unwell to engage in advance care planning. (Most would be too unwell for transfer even if they wanted this). There has been a decrease in less unwell cancer/and non-COVID, non-cancer referrals where advance care planning may be more possible.’* [Hospital Palliative Care Team, Adult Service, England]

Because of this, many respondents spoke about how ACP conversations felt rushed and forced, rather than spending the necessary time needed to adopt a holistic and person-centered approach to discussions:

> *‘ACP was needed to be done quickly and it wasn’t always done at the right time, right place by the right person.’* [Hospital Palliative Care Team, Adult Service, Scotland]

#### Between teams and services level

##### Theme 6: Sharing ACP information

A pre-existing challenge exacerbated by COVID was the sharing of ACP information between services. Different services often used different electronic systems that did not allow for seamless transfer of patient ACP information:

> *‘The ability to share information between primary and secondary care, out-of-hours services and a mixture of [Local Authority] and privately owned care homes.’* [Multiple Settings, Adult Service, Wales]

#### Section 2: Changes to support ACP

This section presents two themes and five sub-themes representing the changes that services made to support ACP during the pandemic.

##### Theme 1: Adapting local processes

###### Sub-theme 1: Prioritisation of escalation planning and DNACPR conversations

One adaptation was to prioritise certain components of ACP (such as treatment escalation plans, Do Not Attempt Cardio-pulmonary Resuscitation, Recommended Summary Plan for Emergency Care and Treatment forms) felt to be of particular importance during the pandemic:

> *‘Frailty nurses have been involved in ensuring that [many] residents in residential care in [locality] have an ACP & treatment escalation plan. Historically ACP for patients known to hospice is high. However, we are ensuring that all patients on IPU & community [register] have treatment escalation plans.’* [Multiple Settings, Adult Service, England]

###### Sub-theme 2: Normalisation and integration of ACP

Another adaptation to local processes was an explicit effort made by services to integrate and embed ACP discussions into everyday clinical practice. This meant proactively initiating, reviewing, and updating ACPs for all COVID and non-COVID patients, alongside ensuring that ACP discussions were routinely reviewed in multidisciplinary team meetings:

> *‘Actively reviewing the outpatient caseload and community caseloads and targeting people without an ACP and broaching this with them more robustly.’* [Multiple Settings, Adult Service, England]

Respondents reflected on the pragmatic and practical steps taken, including having conversations earlier and integrating discussions as a routine practice that was completed on patient referral, admission, and discharge:

> *‘Routinely including option of ACP for all new referrals. Completing treatment escalation planning forms for patients in the community and on discharge from the hospice.’* [Multiple Settings, Adult Service, England]

##### Theme 2: Adapting local structures

###### Sub-theme 3: Using technology to support ACP

One structural change that was made to support ACP discussions was the use of technology. Despite the challenges reported on having virtual and telephone ACP discussions, many respondents reflected on how using these technologies as a form of communication was a way in which they adapted to the pandemic:

> *‘Doing more ACP over the telephone which staff have had to adapt to doing. Patients are understanding the need of social distancing and the impact of COVID - 19.’* [Multiple Settings, Adult Service, England]

Services also used technology to support ACP by refining information technology systems. Predominantly, this included the implementation and documenting of ACP on patients’ electronic record and/or adapting electronic forms so that they were COVID-specific:

> *‘We have had ACP discussions on the phone and via video consultations, we have completed ‘paper’ ACP documents electronically.’* [Home Palliative Care Team, Adult Service, England]
>
> *‘[Name of system] was used where possible which was a new electronic way of recording ACP discussions that had just been finalised for use.’* [Hospital Palliative Care Team, Adult Service, Scotland]

###### Sub-theme 4: Shifting resources

Some respondents reported shifting resources between services as a means to adapt to COVID-19, including increased ACP demands. This was usually through delegating certain staff members with the specific responsibility of taking an active role in supporting ACP discussions:

> *‘Clinical nurse specialist team taking on a much more active role in supporting these conversations.’* [Multiple Settings, Adult Service, England]
>
> *‘Much work from the day hospice team supporting people who have had a GP letter about DNACPR and wish to discuss it further.’* [Inpatient Palliative Care Unit, Adult Service, England]

###### Sub-theme 5: Adapting fast through collaboration

A common change that services made to support ACP during COVID was establishing new, or developing already-existing, networks of support and integrated working within and between teams and services. A heavy emphasis was reported on using these networks to adapt fast through collaboration, usually by drawing on the knowledge and skills of specialists in palliative care who were experienced in ACP. The networks formed and types of collaboration that occurred were considerable. An overview of these collaborative changes with quotes can be seen in Figure 2.

**Figure 2:**
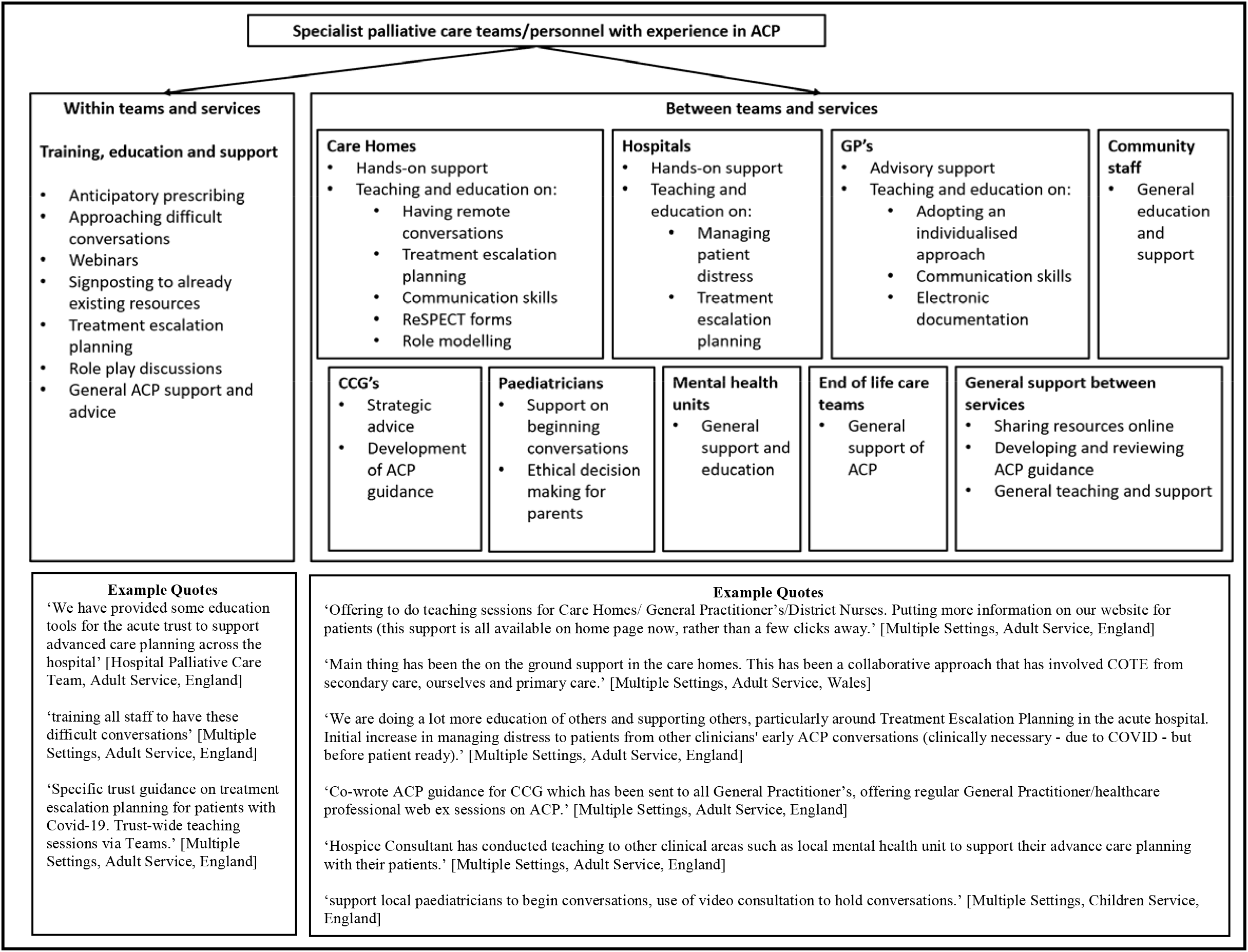
An overview (with example quotes) of the collaboration networks that were established and developed during COVID-19 and how these were used to support ACP

## Discussion

Using the Social Ecological Model, our findings demonstrate how the COVID-19 pandemic exacerbated already-existing challenges to conducting high-quality and timely ACP. At the individual level, the main challenge was maintaining an individualised approach ^(13)^ and making complex decisions in the face of extreme clinical uncertainty ^(13, 14, 28)^. At the within- and between-teams level, racing against rapid deterioration ^(29-31)^ and sharing of ACP-related information ^(13, 32)^ were reported as challenging. Though clinical uncertainty about COVID-19 had similarities to other critical illnesses, ^(32)^ the depth of uncertainty in a disease of which almost nothing was known was of a different order in this pandemic, bringing unique challenges to ACP.

This study shows how COVID-specific challenges made delivering high quality ACP difficult. These occurred at individual (limiting choices of place of care/death), interpersonal (COVID-specific communication difficulties), within-teams (a rapid increase in the volume of ACPs combined with sudden decrease in staff/services) and national (delivering ACP in a national context of fear and uncertainty) levels. The Social Ecological Model illuminated how a national context of fear provided a contextual backdrop through which the various challenges are better understood. These worries may be viewed through the ‘four horsemen of fear’ concept ^(33)^ in which COVID-19 precipitated bodily, interpersonal, cognitive, and behavioural fears. These fears were brought into ACP conversations by patients, and their families, and health professionals, disrupting their ability to engage in ACP conversations as effectively as they would have liked.

In adapting to these challenges, services made changes to structures and processes of care. There is already evidence of the benefits of some of these, such as having earlier ACP discussions^(13, 34)^ and training aimed at facilitating healthcare professionals’ skills/confidence in communicating ACPs. ^(35-37)^ Recent work has also demonstrated the feasibility and effectiveness of having virtual discussions with patients/families during COVID-19, ^(38, 39)^ and resources have been developed to support healthcare professionals to navigate the challenges and sensitivities of virtual difficult conversations. ^(40-42)^

However, some changes induced by the pandemic, such as reducing ACP to specific components were less helpful. This is because ACP is a multi-component process, not a one-time event/document, not least because preferences and priorities may change. ^(6-8, 10, 12, 34, 43-46)^ Delivering ***all*** of the multiple components of ACP, and delivering them well, is important to ensure inclusive, holistic, and individualised care that focuses on what matters most to patients. ^(47)^ Whilst understandable in the pandemic context, emphasis on discrete components of ACP may jeopardise the individualised and holistic qualities essential for the delivery of high quality and comprehensive ACP, and runs the risk of making ACP a ‘tick box exercise focused on a predetermined list of preferences’.^(43)^ This is a concern raised by the public and clinical communities. ^(43, 48, 49)^

### Considerations for clinical practice and policy

COVID-19 has provided an opportunity to re-think ACP in which the starting point to any discussion is always the values and priorities of patients themselves. Initially, these discussions are likely to be broad in nature, with their focus then narrowing in line with the more immediate concerns of individuals. ^(47)^

Some changes to support ACP were temporary and may be dropped post-pandemic (such as shifting of resources and focusing on specific components of ACP), but innovative changes that showed promise may be amplified and sustained. Changes such as learning fast through collaboration, training to support ACP, the integration of ACP into everyday clinical practice, and use of virtual technology are important to maintain as the need for palliative care is estimated to rise considerably ^(18)^ and need for ACP will not be met by specialists alone. ^(50-52)^ In facilitating these changes, Table 4 provides questions for health professionals and policymakers – in the UK and beyond - to consider when conducting ACP during a pandemic and in clinical practice more generally. Most importantly, policymakers in any given country need to consider how high-quality ACP can be resourced as a part of standard care.

**Table 4:**
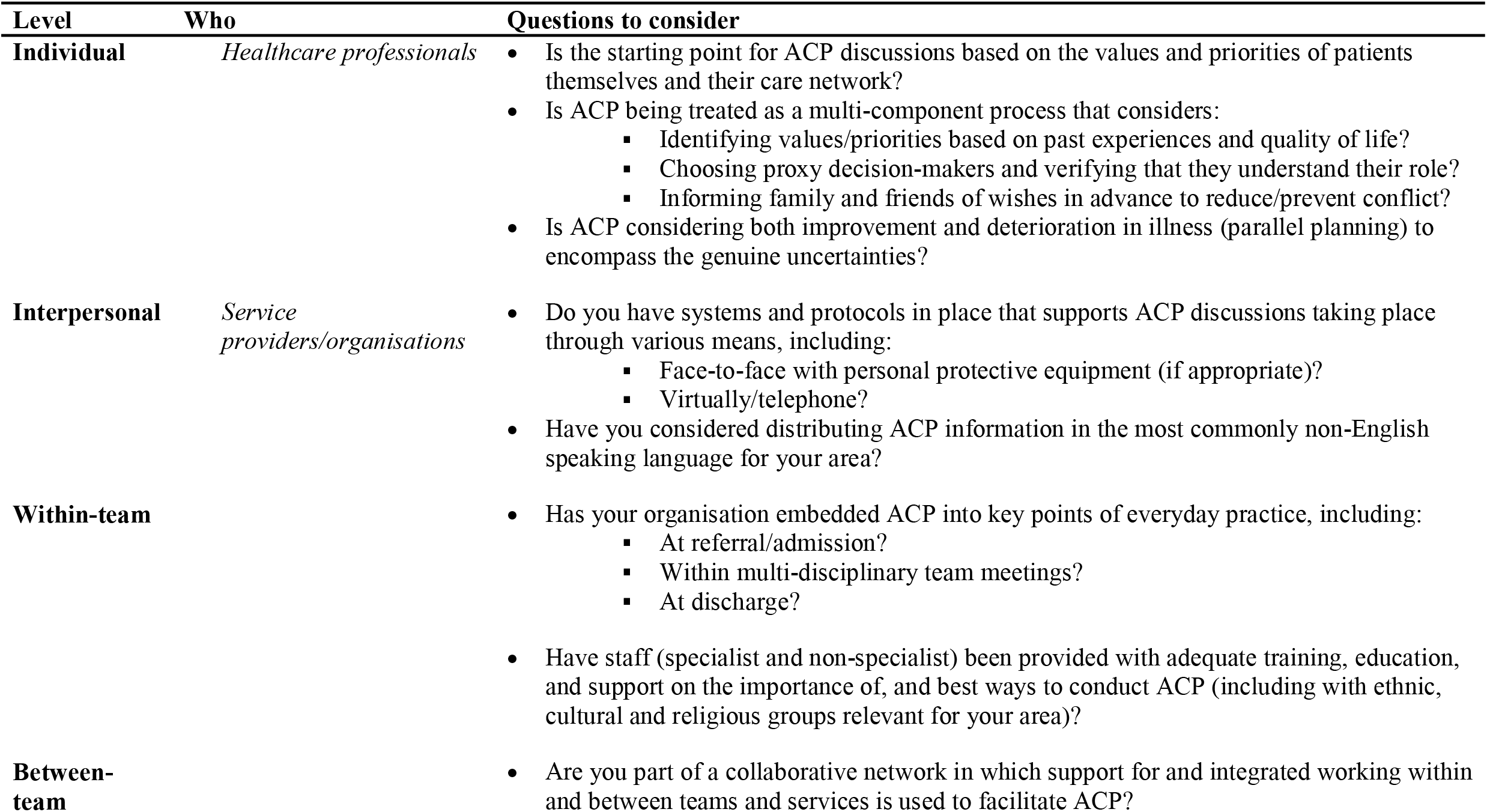
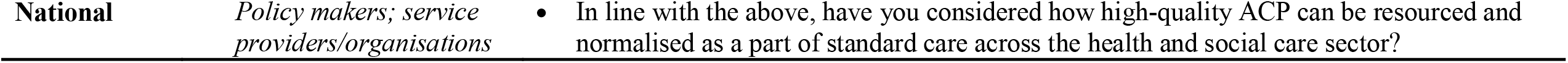
Multi-level considerations for conducting high-quality ACP during a pandemic and clinical practice more generally

| Level | Who | Questions to consider |
| --- | --- | --- |
| **Individual** | *Healthcare professionals* | • Is the starting point for ACP discussions based on the values and priorities of patients themselves and their care network?<br>• Is ACP being treated as a multi-component process that considers:<br>▪ Identifying values/priorities based on past experiences and quality of life?<br>▪ Choosing proxy decision-makers and verifying that they understand their role?<br>▪ Informing family and friends of wishes in advance to reduce/prevent conflict?<br>• Is ACP considering both improvement and deterioration in illness (parallel planning) to encompass the genuine uncertainties? |
| **Interpersonal** | *Service providers/organisations* | • Do you have systems and protocols in place that supports ACP discussions taking place through various means, including:<br>▪ Face-to-face with personal protective equipment (if appropriate)?<br>▪ Virtually/telephone?<br>• Have you considered distributing ACP information in the most commonly non-English speaking language for your area? |
| **Within-team** |  | • Has your organisation embedded ACP into key points of everyday practice, including:<br>▪ At referral/admission?<br>▪ Within multi-disciplinary team meetings?<br>▪ At discharge?<br>• Have staff (specialist and non-specialist) been provided with adequate training, education, and support on the importance of, and best ways to conduct ACP (including with ethnic, cultural and religious groups relevant for your area)? |
| **Between-team** |  | • Are you part of a collaborative network in which support for and integrated working within and between teams and services is used to facilitate ACP? |

### Strengths, limitations, and future research

This survey received a high number of respondents from all palliative care settings: Hospice UK report approximately 200 hospice services across the UK; 168 (∼ 84%) responded to this survey. ^(53)^ The timely delivery of the survey enabled capture of changes across the peak of the first wave of COVID-19 in the UK.

ACP is influenced and moderated by contextual and cultural-dependant factors. ^(54, 55)^ Whilst many of the findings of this paper may be applicable in these contexts, more research that explores international and cultural differences regarding ACP during COVID-19 is needed. Survey data was collected at a single time-point and so the processes through which challenges to ACP changed over time, and the longer-term impact, sustainability, and effectiveness of changes are not always clear.

## Conclusion

Many challenges to providing high quality ACP during COVID-19 pre-dated the pandemic, whilst others were COVID-19 specific, or markedly exacerbated by the pandemic. Professionals and healthcare providers need to ensure ACP is well-founded for individuals, and genuinely tailored to their values and priorities, and attuned to their ethnic, cultural and religious context. Policymakers for health and social care need to consider carefully how high-quality ACP can be resourced and normalised as a part of standard health-care ahead of future pandemic waves.

## Data Availability

Applications for use of the survey data can be made for up to 10 years, and will be considered on a case by case basis on receipt of a methodologically sound proposal to achieve aims in line with the original protocol. The study protocol is available on request. All requests for data access should be addressed to the Chief Investigator via the details on the CovPall website (https://www.kcl.ac.uk/cicelysaunders/research/evaluating/covpall-study, and) and will be reviewed by the Study Steering Group.

## Author Contributions

IJH is the grant holder and chief investigator; KES, MM, FEM, CW, NP, LKF, SB, MBH and AO are co-applicants for funding. IJH and CW, with critical input from all authors, wrote the protocol for the CovPall study. MBH, AO, and RC co-ordinated data collection and liaised with centres, with input from IJH. AB, FEM, and LKF analysed the data. All authors had access to all study data, discussed the interpretation of findings and take responsibility for data integrity and analysis. AB, FEM, and LKF drafted the manuscript. All authors contributed to the analysis plan and provided critical revision of the manuscript for important intellectual content. IJH is the guarantor. Sites who contributed to this work can be found in supplementary file 3.

## The CovPall study group

CovPall Study Team: Professor Irene J Higginson (Chief Investigator), Dr Sabrina Bajwah (Co-I), Dr Matthew Maddocks (Co-I), Professor Fliss Murtagh (Co-I), Professor Nancy Preston (Co-I), Dr Katherine E Sleeman (Co-I), Professor Catherine Walshe (Co-I), Professor Lorna K Fraser (Co-I), Dr Mevhibe B Hocaoglu (Co-I), Dr Adejoke Oluyase (Co-I), Dr Andrew Bradshaw, Lesley Dunleavy and Rachel L Cripps.

CovPall Study Partners: Hospice UK, Marie Curie, Sue Ryder, Palliative Outcome Scale Team, European Association of Palliative Care (EAPC), Together for Short Lives and Scottish Partnership for Palliative Care.

## Acknowledgments

This study was part of CovPall, a multi-national study, supported by the Medical Research Council, National Institute for Health Research Applied Research Collaboration South London and Cicely Saunders International. We thank all collaborators and advisors. We thank all participants, partners, PPI members and our Study Steering Group. We gratefully acknowledge technical assistance from the Precision Health Informatics Data Lab group (https://phidatalab.org) at National Institute for Health Research (NIHR) Biomedical Research Centre at South London and Maudsley NHS Foundation Trust and King’s College London for the use of REDCap for data capture.

## CovPall Data Sharing Statement

Applications for use of the survey data can be made for up to 10 years, and will be considered on a case by case basis on receipt of a methodological sound proposal to achieve aims in line with the original protocol. The study protocol is available on request. All requests for data access should be addressed to the Chief Investigator via the details on the CovPall website (https://www.kcl.ac.uk/cicelysaunders/research/evaluating/covpall-study, and) and will be reviewed by the Study Steering Group.

