## Supplementary File 2 for "Understanding and addressing challenges for Advance Care Planning in the COVID-19 pandemic: An analysis of the UK CovPall survey data from specialist palliative care services"

**Supplementary file 2: Procedures for CovPall survey**

Services were identified and contacted through national and multinational gatekeeper palliative care and hospice organisations (see acknowledgements and supplementary file 3 for details) and asked that their medical or nursing lead, or their nominee, complete an on-line survey available via a link. The email attached the participant information sheet with details of the study rationale, ethical approval, data protection and management, investigators and contacts for further information or concern. No incentives were offered for completion. Completion was taken to indicate informed consent. The CovPall study was presented at relevant on line meetings and discussed with gatekeeper organisations and in blogs, to inform the methods, questions and to raise awareness and engagement.

We developed and piloted a secure, password-protected web-based data entry portal (in the Research Electronic Data Capture (REDCap) at the study coordinating centre, King’s College London. Services could keep their identity hidden if they wished, but most chose to provide an email for contact. Data were anonymised before analysis.

The questionnaire was developed and piloted by the CovPall study team building on an earlier survey of Italian hospices, adding questions on the impact of and response to COVID-19. It was intended to be brief, taking around 30 minutes to complete as we recognised staff were busy, and comprised seven sections: region and responder; PC/hospice management and services offered pre-COVID-19; experiences of patients and staff with COVID-19; changes to services overall; changes in specific settings; shortages experienced; symptom management. Free-text explanatory comments were invited in all sections and respondents also indicated whether and how they could be contacted. Respondents could save and complete the questionnaire later if they wished. The questionnaire is available in supplementary file 1.

The survey opened on April 23rd and closed July 31st 2020. The study coordinating centre replied actively to respondents’ queries or requests for help.

In most cases, data collection and entry was done electronically via the REDCap link directly by the services without problems, however a few palliative care services had difficulties accessing the online survey due to local computer procedures. In these instances, they were sent the survey in paper format, completed this and returned it to King’s to enter this data, or were interviewed over the phone and data entry sheet was completed by the researchers. The research team also audited the data weekly to ensure data entry completeness and sent monthly missing data and incomplete entry reports to the research associates and administrators, and where consent permitted to relevant respondents to check validity.
