## Supplementary File 3 for "Understanding and addressing challenges for Advance Care Planning in the COVID-19 pandemic: An analysis of the UK CovPall survey data from specialist palliative care services"

We would like to thank all the palliative care services and leads across the world for responding to the CovPall online survey.

The following persons and services indicated they were happy to be acknowledged for responding to the CovPall online survey.

Alexander Devine Children’s Hospice Service

Alison Wardrop District Nursing Sister, Hywel Dda University Health Board

Ashgate Hospicecare

Bangalore Baptist Hospital Palliative Care Service

BCUHB West HSPCT

Beatson West of Scotland Cancer Centre HSPCT

Beaumond House Community Hospice

Betsi Cadwaladr University Health Board SPCT

Birmingham St Mary’s Hospice

Bluebell Wood Children's Hospice

Bolton NHS Foundation Trust Specialist Palliative Care Team

Bury Community Specialist Palliative Care Team

Cambridge University Hospital NHS Foundation Trust Palliative Care Service

Carmarthenshire Specialist Palliative Care Service, Hywel Dda University Health Board

Center for Palliative Care, University Hospital Cologne, Germany

Cesta domů, home hospice

Chelsea and Westminster Hospital

CHFT Hospital SPCT

Children’s hospices across Scotland (CHAS)

City Hospice Cardiff

Claire House Children's Hospice

Clinique de Médecine Palliative, CHU de Lille, France

CNWL UCLH

Community based clinical nurse specialists

Community Specialist Palliative Care Team, St Ann's Hospice, Salford

Consultant Dr Paul Coulter, CNS Elizabeth Anderson and CNS Kellyann O'Neill, Palliative Care Team, Inverclyde Royal Hospital

Cornwall Hospice Care

Croydon Health Services Macmillan Specialist Palliative Care Team

Cwm Taf Morgannwg SPCT North

Department of Palliative Care, Homerton University Hospital NHS Foundation Trust

Department of Palliative care, Sheffield Teaching Hospitals NHS Foundation Trust

Department of Palliative Medicine, LMU Munich, Germany

Department of Palliative Medicine, Southern Sector, South East Sydney Local Health District, New South Wales, Australia

Deutsche PalliativStiftung

Dorothy House Hospice

Dove House Hospice, Hull

Dr Charles Daniels, Medical Director, St Luke’s Hospice Harrow and Consultant in Palliative Medicine, LNWHUT.

Dr Jonathan Downie, Consultant in Paediatric Palliative Medicine, Supportive and Palliative Care Team, Royal Hospital for Children, Glasgow

East Anglia's Children's Hospices (EACH)

East Cheshire Hospice

East Sussex Healthcare NHS Trust Supportive and Palliative Care Team

Eastern Health

Ed Dubland MD CCFP pc

Ellenor Adult Services

Ellenor Children’s Services

Ellenor Hospice, Coldharbour Road, Gravesend, Kent

Ellenor Inpatient Ward

Ellenor Wellbeing Service

Elvis J Miti for Community Palliative Care, UZIMA Project Ndanda, Mtwara, Tanzania.

ESNEFT specialist palliative care team

Family Support Team, Children's Hospices Across Scotland

Farleigh Hospice

Felicia Kontopidis, Journey Home Hospice

Fondazione Antea, Rome, Italy

Fondazione FARO, Turin, ITALY

Forest Holme Hospice, Poole Hospital NHS Foundation Trust

Forget Me Not Children's Hospice

Garden House Hospice Care

GHNHSFT Specialist Palliative Care Community team

Glasgow Royal Infirmary Hospital Specialist Palliative Care Team

Greenwich & Bexley Community Hospice

GSTT Community Palliative Care Team

Hambleton & Richmondshire SPCT

Hampshire Hospitals NHS Foundation Trust

Harrogate and District NHS Foundation Trust Palliative Care Team

Heart of Kent Hospice

Helen & Douglas House

Hospice De Wingerd (Charim Care Group)

Hospice Isle of Man

Hospice of St. Francis Berkhamsted, Herts

Hospice of The Good Shepherd, Chester

Hospice Waikato

Hospiscare Devon

Hospital Divina Providencia, El Salvador

Hospital Palliative Care Team, Kingston Hospital NHS Foundation Trust

Hospital Palliative Care Team, University Hospitals of North Midlands

Hospital Palliative Care team, University Hospital Southampton

Hospital Selayang Palliative Care Unit

Hospital Specialist Palliative care team

Hospital Specialist Palliative Care Team, University Hospitals of Leicester NHS Trust

Hull University Teaching Hospitals NHS Trust

Instituto de Investigaciones Médicas Alfredo Lanari, Universidad de Buenos Aires

Isabel Hospice

J Yeomans, St Richards Hospice (Worcester)

James Cook University Hospital Acute SPCT

Thitima Phosri

John Taylor Hospice

Helen Hubert, Palliative care and End of Life Care Pharmacist, St Richard’s Palliative Care Team

Katharine House Hospice Banbury

Kauniala Hospital

Kenelm F McCormick M.D., Akron, Ohio

Kilbryde Hospice, South Lanarkshire

Kim Jones, Deputy Head of Clinical Services at Hospice of the Valleys

King's College Hospital NHS Foundation Trust

Klinik Susenberg, Zürich

Knysna Sedgefield Hospice

Lanarkshire SPC service

Laois Offaly Palliative Care Service Cloneygowan Co Offaly

Leeds Teaching Hospitals NHS Trust Palliative Care Team

Limewood Dementia Service, Stafford

London Northwest Healthcare University Trust

LOROS Hospice, Leicester

Lynn Bushor, Department of Veteran's Affairs

Macmillan Unit / Royal Bournemouth & Christchurch

Manchester Foundation Trust - MRI

Manchester Foundation Trust WTWA

Maricruz Macias Montero, Hospital General de Segovia, Seccion de Geriatria.

Marie Curie Community Nursing Service London

Marie Curie Hospice, Cardiff and the Vale

Marie Curie Hospice West Midlands

Marie Curie Hospice, Hampstead

Marie Curie Services in Lothian

Martlets Hospice

Martlets Hospice, Brighton

Maura Farrell Miller, Director, Hospice and Palliative Care Program, Department of Veterans Affairs Medical Center, Florida

Mellannorrlands Hospice Sundsvall Sweden

Michael Sobell Hospice and Harlington Hospice

Mid Cheshire Hospital NHS Trust Specialist Palliative Care Team

Midhurst Macmillan Service (Sussex Community NHS Foundation Trust)

Municipal Hospital Dr Cornel Igna, Palliative Care Department from Campia Turzii

Myton Hospice

Newham University Hospital Specialist Palliative Care Team - Barts Health NHS Trust

NHS Ayrshire & Arran Supportive Care Team

NHS Fife Specialist Palliative Care Service

NHS Grampian Specialist Palliative Care Team

NHS Specialist Palliative Care Unit

Nightingale House Hospice, North Wales

Ninewells Hospital Palliative Care Team

North London Hospice

Northern Ireland Hospice Adult Service

Northumbria Healthcare NHS Foundation Trust

Nottingham University Hospitals NHS Trust

Ofra Fried, Palliative Care Specialist, Townsville University Hospital, Queensland

Overgate Hospice

Palliativa Care Unit. west health Area. Valladolid. Castilla y León .Spain

Palliative and End of Life care Team, Newcastle upon Tyne Hospitals NHS Foundation Trust

Palliative Care Department, Bangalore Baptist Hospital

Palliative Care Service, Chaim Sheba Medical Center, Ramat-Gan, Israel

Palliative Care Service, Gloucestershire Hospitals NHS Foundation Trust

Palliative Care Team-Bassett Medical Center, Cooperstown NY

Palliative Care Team, Centre for Pain Management and Palliative Care, Haukeland University Hospital, Bergen, Norway

Palliative Care Unit, Bolzano, Italy

Palliative Care Unit, Bassini Hospital, Cinisello Balsamo, Milan, Italy

Palliative Care, Kantonsspital Olten, Switzerland

Palliative Medicine, UHDB NHS Trust

Palliaviva

Peace Hospice Care

Pembrokeshire Specialist Palliative Care service, Hywel Dda University Health Board

Phyllis Tuckwell Hospice Care

Pilgrims Hospices in East Kent

Pippa Hawley, Medical Director, Pain and Symptom Management Palliative Care, BC Cancer.

Portsmouth Hospital NHS Trust Palliative Care Service

Princess Alice Hospice

Prof Olaitan A Soyannwo

Queen Elizabeth University Hospital Glasgow Specialist Palliative Care Team, and Prince and Princess of Wales Hospice Glasgow

Queenscourt Supportive & Specialist Palliative Care Service

Rotherham NHS Foundation Trust Palliative Care Team

Rowans Hospice

Rowcroft Hospice

Royal Berkshire Foundation Trust Hospital Palliative Care Team

Royal Trinity Hospice

Sabar Health Home Hospital

Saint Francis Hospice

Salford Royal Hospital Specialist Palliative Care Team

Salisbury Specialist Palliative Care service

SAPV des OSP Karlsruhe

SCCU SEVILLA SUR, SAS. SPAIN.

Sengkang General Hospital, Singapore

Shooting Star Children's Hospice

Sobell House, Oxford University Hospitals

South East Palliative Care Services

South East Palliative Care, University Hospital Waterford, Ireland

South Tees Community Specialist Palliative Care MRC

South Tipperary Hospice Homecare Team

South Tyneside District Hospital Specialist Palliative Care Team, South Tyneside and Sunderland NHS Foundation Trust

Specialist Palliative Care Hospital Team. East Kent University Foundation Trust

Specialist Palliative Care Service South Eastern Health and Social Care Trust

Specialist Palliative Care Team Imperial College Healthcare NHS Trust

Specialist Palliative Care Team, Coventry and Warwickshire Partnership NHS Trust

Specialist Palliative Care Team, West Middlesex University Hospital, Chelsea and Westminster NHSFT

Specialist Palliative Care Service, County Durham and Darlington NHS Foundation Trust

St Ann's Hospice

St Barnabas Hospice, Lincolnshire

St Barnabas House, Worthing

St Bartholomew’s Hospital, Barts Health

St Catherine's Hospice, Preston

St Christopher's

St Clare Hospice, Essex

St Columba's Hospice, Edinburgh

St Gemma's Hospice - Leeds, West Yorkshire.

St Helena

St John of God Murdoch Hospital

St John's Hospice, Doncaster, RDASH

St Johns Hospice, Lancaster

St Leonard's Hospice, York

St Luke's Cheshire Hospice

St Luke's Combined Hospices, Cape Town, South Africa

St Luke's Hospice, Sheffield.

St Luke’s Hospice, Kenton, Harrow covering London boroughs of Brent and Harrow. Ursula Reeve, Director of Patient Services. Cathy Hanrott, Hospice Service Navigator

St Margaret’s Hospice, Somerset.

St Martha's Hospital Palliative Care Team

St Mary's Hospice, Ulverston, Cumbria

St Michael's Hospice, Hastings and Rother

St Oswald's Hospice, Newcastle

St Peter's Hospice

St Richards Hospice

St Wilfrid's Hospice, Chichester

St Wilfrid's Hospice, Eastbourne.

St. Luke's Hospice Community Services South West Essex

Stockport Specialist Palliative Care Service

Strathcarron Hospice

Sue Ryder Leckhampton Court Hospice

Sue Ryder Manorlands Hospice

Susanna Sandöy, Capio Asih Nacka,

Swansea Bay University Health Board Specialist Palliative Care Service (which includes Ty Olwen Hospice)

Symptom Control and Palliative Care Team, Royal Marsden NHS Foundation Trust

Tatiana Chavouzi

Tayside Palliative Care Service

The Arthur Rank Hospice, Cambridge

The Camden, Islington ELiPSe Palliative Care Team

The Hillingdon Hospital Palliative Care team

The Prince of Wales Hospice, Pontefract

The Wisdom Hospice and Medway Community Healthcare

TOPAT Zuyderland MC

Trinity Hospice & Brian House Children's Hospice

Trinity Hospice, Blackpool and Blackpool Teaching Hospitals

Tynedale Hospice at Home

Unità di Cure Palliative - Hospice Casale Monferrato ASL AL Piemonte

University Hospital Lewisham Macmillan Specialist Palliative Care Team, Lewisham and Greenwich NHS Trust

University Teaching Hospital Lusaka Adult Medicine Palliative Care Unit

Waterford Hospice Homecare Team

Weldmar Hospicecare

West Suffolk Hospital NHS Foundation Trust

Western Australia Paediatric & Adolescent Palliative Oncology, Perth Children's Hospital

Weston Hospicecare

Whittington Health Palliative Care Service

Wigan & Leigh Hospice

Wrighinton, Wigan and Leigh NHS Teaching Trust (Supportive and Palliative Care Team)

York Teaching Hospital Foundation Trust

York Teaching Hospitals

Zuercher Lighthouse

Zuzana Kremenova
